# Prevalence of Metabolic Syndrome in the United States National Health and Nutrition Examination Survey (NHANES) 2011-2018

**DOI:** 10.1101/2021.04.21.21255850

**Authors:** Xiaopeng Liang, Benjamin Or, Man Fung Tsoi, Ching Lung Cheung, Bernard MY Cheung

**Author notes:** **Correspondence** Prof. Bernard M.Y. Cheung, PhD, FRCP, Department of Medicine, Queen Mary Hospital, 102 Pokfulam Road, Hong Kong.

## Abstract

**Importance:** Metabolic syndrome (MetS) is a cluster of risk factors presaging the development of cardiovascular diseases and diabetes. It is a risk factor for severe coronavirus disease 2019 (COVID-19).

**Objective:** To estimate the prevalence of MetS in the US National Health and Nutrition Examination Survey (NHANES) 2011-2018.

**Design, Setting, and Participants:** This cohort study included 22370 eligible participants aged ≥20 years from the NHANES 2011-2018.

**Main Outcome and Measure:** MetS was defined as the presence of at least three of these components: central obesity, reduced high-density lipoprotein, elevated triglycerides, elevated blood pressure and elevated fasting blood glucose. The prevalence of MetS was estimated taking into account the complex sampling. The time trend was evaluated using logistic regression. Annual percentage changes (APC) were used to measure the trends in MetS prevalence.

**Results:** The prevalence of MetS was 36.2% (95% CI, 32.3-40.3), 34.8% (95% CI, 32.3-37.4), 39.9% (95% CI, 36.6-43.2) and 38.3% (95% CI, 35.3-41.3) in 2011-2, 2013-4, 2015-6, 2017-8, respectively (*P* for trend =.08). Among the MetS components, the prevalence of elevated glucose increased from 48.7% (95% CI, 45.9-51.5) in 2011-2 to 64.3% (95% CI, 61.0-67.4) in 2017-8 [*P* for trend <.001; APC=11.7, (95% CI, 3.5-21.0)]. The prevalence of MetS in non-Hispanic Asian increased from 21.8% (95% CI, 16.7-28.0) in 2011-2 to 31.2% (95% CI, 27.4-35.3) in 2017-8 [*P* for trend <.001; APC=14.6, (95% CI, 2.5-34.8)].

**Conclusion and Relevance:** The prevalence of MetS remained stable from 2011 to 2018, but increased among non-Hispanic Asians. Lifestyle modification is needed to prevent metabolic syndrome and the associated risks of diabetes and cardiovascular disease.

## Background

Metabolic syndrome (MetS) is a cluster of conditions comprising central obesity, elevated blood pressure, hyperglycemia, hypertriglyceridemia and reduced high-density lipoprotein-cholesterol (HDL) that are associated with the development of type 2 diabetes mellitus and cardiovascular disease.^1^ In the National Cholesterol Education Program’s Adult Treatment Panel III definition, a person with three or more criteria above is deemed to have MetS.^2^ We and other groups have shown that MetS is also associated with hypertension, stroke, cancer and increased mortality.^3-5^ Recent studies have revealed that MetS and its components are highly associated with the susceptibility to severe acute respiratory syndrome coronavirus 2 (SARS-CoV-2) infection and the severity of coronavirus disease 2019 (COVID-19).^6,7^ Therefore, diagnosing metabolic syndrome is clinically important because it prompts clinicians to look for the myriad of diseases associated with it, helps patients to understand the causes of their seemingly diverse ailments, and paves the way for the prevention of the complications of MetS, such as cardiovascular diseases, and severity of COVID-19, through pharmacologic and non-pharmacologic therapy.^8^

At the societal level, the prevalence of MetS is an index of the cardiometabolic health of the population. In the US, it has been reported many times that the prevalence of diabetes has been increasing and has reached epidemic proportions.^9^ Abdominal obesity, or central obesity, leads to a state of systemic inflammation and insulin resistance. It plays a major role in the pathogenesis of MetS.^10^ Previous reports have highlighted a steady increase in the prevalence of MetS in the US.^11, 12^ However, its prevalence appears to have reached a plateau in 2016, with no evidence of further increases.^13^ The prevalence was 34%, 33%, and 34.7% in 1999-2006,^14^ 2003-2012,^15^ and 2011-2016,^13^ respectively. This may seem surprising against the background of an obesity and diabetes epidemic, which would be expected to increase the prevalence of MetS as they are components of the syndrome.^16^

Socioeconomic status (SES), reflecting education, occupation and income, is known to be a powerful predictor of morbidity and mortality.^17^ Previous studies have shown greater decline in the prevalence of obesity, diabetes and cardiovascular diseases among people of higher SES in the US.^18^ The prevalence and time trends in MetS in people with different SES warrants investigation.

Therefore, we used the latest figures up to 2018 to estimate the latest prevalence of MetS in US adults, and in gender, age, ethnic and socioeconomic subgroups.

## Methods

### Description of the study

The US National Health and Nutrition Examination Survey (NHANES) is a continuous and longitudinal survey since 1999. It received ethical approval from the National Center for Health Statistics Ethics Review Board. Written consent was obtained from all adult subjects. A cross-sectional, stratified, multistage probability sampling method was used to obtain a nationally representative sample of the US population ≥ 20 years of age. Demographics, examination, questionnaire, and laboratory data covering the period 2011-2018 were extracted.^19^ Pregnant women were excluded to reduce bias associated with weight, hypertension, and gestational diabetes.

### Definition

Self-reported race/ethnicity was categorized into the following subgroups: Hispanic, non-Hispanic white, non-Hispanic black, non-Hispanic Asian, and ‘other’. Hispanic race/ethnicity includes Mexican American and other Hispanic races. Other race/ethnicity includes other non-Hispanic races, including non-Hispanic multiracial. Educational level was categorized into four subgroups: less than 11th grade, high school, some college, and college or above. The household income is based on a comparison of family income with the poverty threshold determined by the U.S. Bureau of Census. The household income was classified as < 130%, 130-349%, and ≥ 350%. MetS was defined according to the National Cholesterol Education Program’s Adult Treatment Panel III as having at least three of the following diagnostic criteria: waist circumference greater than 102 cm in men or 88 cm in women, triglyceride level greater than 150 mg/dL, high-density lipoprotein cholesterol less than 40 mg/dL in men or less than 50 mg/dL in women, systolic blood pressure at least 130 mm Hg or diastolic blood pressure at least 85 mm Hg or taking hypertension medications, or fasting plasma glucose level at least 100 mg/dL or taking diabetes medications.^2, 20^

### Statistical analysis

Statistical analysis was performed in STATA (version 15.0). Four discrete 2-year cycles of continuous NHANES (2011-2012, 2013-2014, 2014-2016, 2017-2018) were analyzed using complex sample weights accounting for unequal probabilities of selection, nonresponse bias, and oversampling. Chi-square tests and Kruskal-Wallis were used to compare categorical and continuous variables, respectively. Trends were assessed using logistic regression after regressing metabolic syndrome on year (prevalence was treated as an outcome variable and cycle year was modeled as a continuous predictor). Annual percentage changes (APC) were calculated to measure the trends in MetS during 2011-2018 using the Joinpoint Regression Program, Version 4.9.0.0 (Statistical Research and Applications Branch, National Cancer Institute). All analyses were 2-tailed, and P<0.05 was considered statistically significant.

## Results

### Characteristics of study participants

We included 22370 adult participants [mean age 47.9 years (95% CI, 47.3-48.6), 48.6% (95% CI, 47.5-49.6) men] from the four most recent 2-year cycles of continuous NHANES (2011-2012, 2013-2014, 2014-2016, 2017-2018). The percentage of adults that underwent an examination in these cycles was 69.5%, 68.5%, 58.7%, and 48.8%, respectively.^21^ Their characteristics of participants included in this study by gender, age group, race/ethnicity and socioeconomic status are shown in Table 1. The percentages of each component of MetS among the study participants are shown in Figure 1. The percentage of elevated glucose increased from 48.7% (95% CI, 45.9-51.5) in 2011-2 to 64.3% (95% CI, 61.0-67.4) in 2017-8 [*P* for trend <.001; APC=11.7, (95% CI, 3.5-21.0)], whereas the percentage of reduced HDL, central obesity, elevated triglyceride and blood pressure remained stable in participants from 2011-2014 to 2017-2018 (All *P* for trend >.05).

**Table 1.**
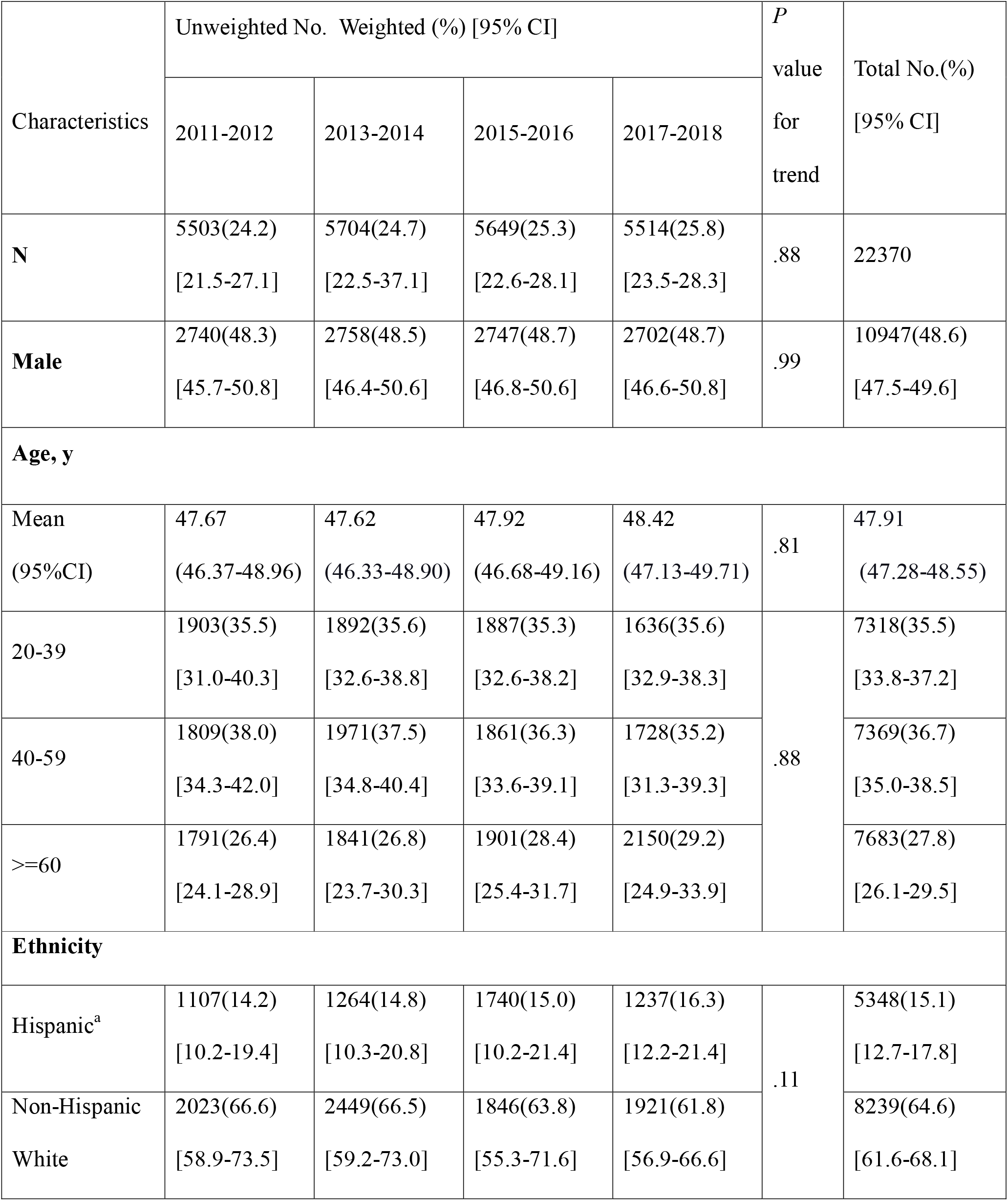
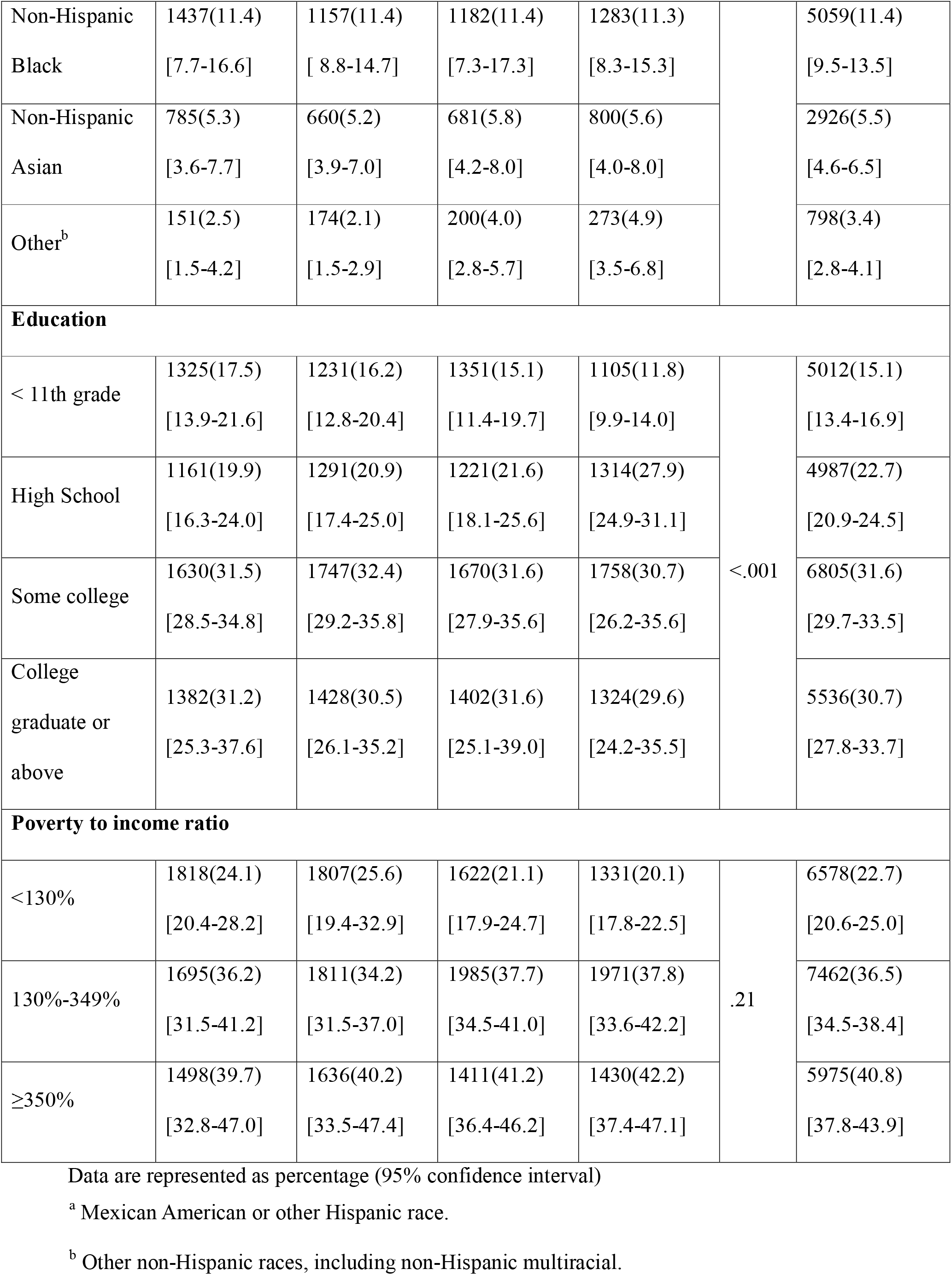
Characteristics of the NHANES Participants Analyzed

**Figure 1.**
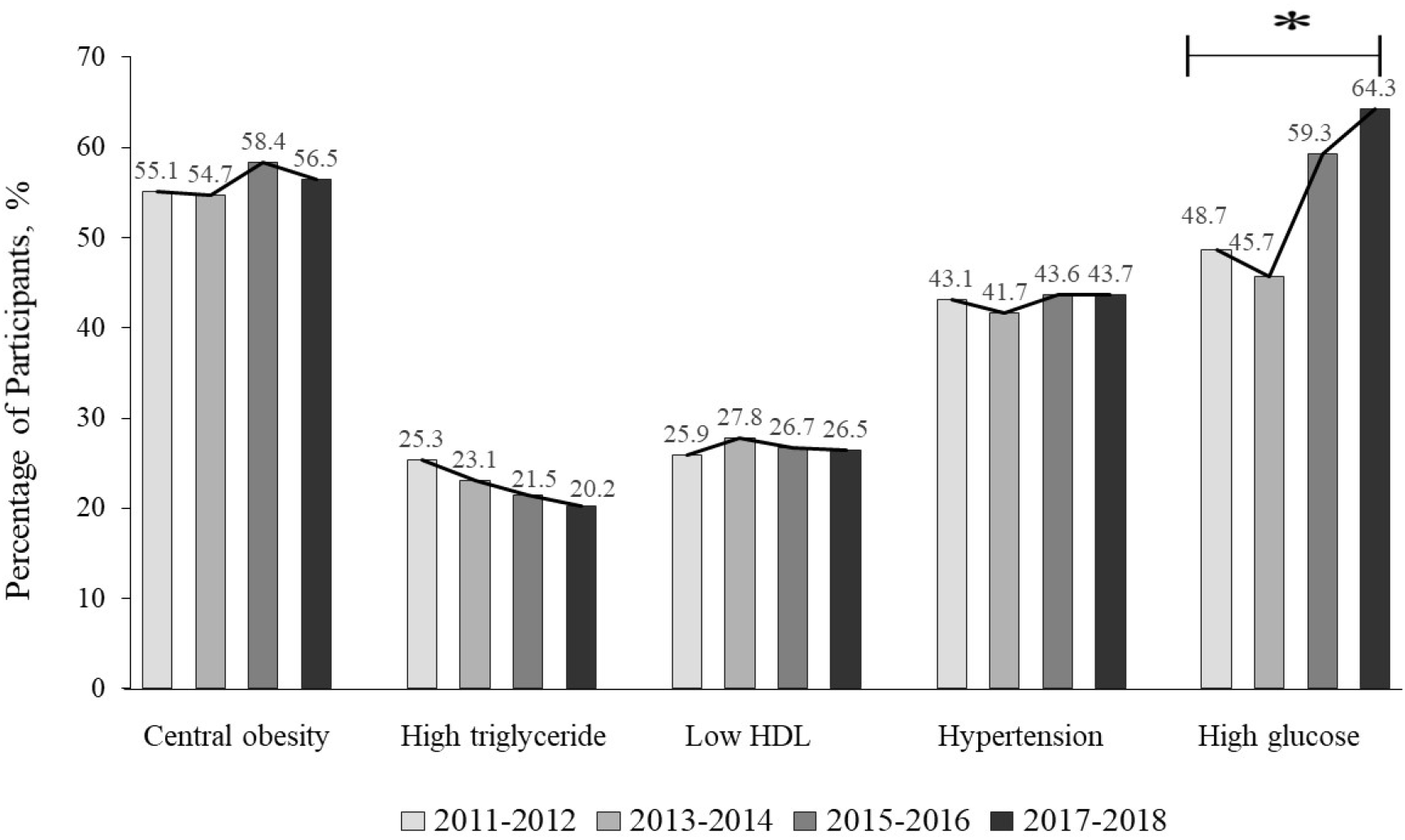
The Percentage of Participants with Each Component of MetS * *P*<.001 for the trend of the MetS components Central obesity was defined as waist circumference ≥102 cm in men or ≥ 88 cm in women. High triglyceride was defined as plasma triglyceride ≥ 150 mg/dL Low HDL was defined as high-density lipoprotein cholesterol (HDL) < 40 mg/dL in men or < 50 mg/dL in women Hypertension was defined as systolic blood pressure ≥ 130 mm Hg, or diastolic blood pressure ≥ 85 mm Hg, or taking hypertension medications High glucose was defined as fasting plasma glucose ≥ 100 mg/dL or taking diabetes medications

### Overall prevalence of MetS in 2011-2018

Among 22370 participants, 5820 fulfilled the criteria for MetS, giving an overall prevalence of 37.3% (95% CI, 35.7-39.0). The prevalence was not significantly different among men and women [37.9% (95% CI, 35.6-40.2) vs 36.8% (95% CI, 34.7-39.0), *P* =.48]. As expected, the prevalence increased with age (Table 2). It increased from 20.1% (95% CI, 18.1-22.4) in the 20-39 age group to 53.7% (95% CI, 50.7-56.7) in those aged ≥ 60 years (*P* <.001). It was highest in non-Hispanic Whites (39.3%; 95% CI, 37.1-41.0) and lowest in non-Hispanic Asians (24.0%; 95% CI, 21.6-26.6).

**Table 2.** Prevalence of Metabolic syndrome by Gender, Age Group, Race/Ethnicity

|  | Unweighted No. (Weighted %) [95% CI] |  |  |  | *P* value<br>for trend<br>cross the<br>years | Total<br>unweighted<br>No.<br>(weighted %<br>)<br>[95% CI] |
| --- | --- | --- | --- | --- | --- | --- |
|  | 2011-2012 | 2013-2014 | 2015-2016 | 2017-2018 |  |  |
| MetS | 1330(36.2)<br>[32.3-40.3] | 1432(34.8)<br>[32.3-37.4] | 1543(39.9)<br>[36.6-43.2] | 1515(38.3)<br>[35.3-41.3] | .08 | 5820(37.3)<br>[35.7-39.0] |
| **Gender** |  |  |  |  |  |  |
| Male | 623(37.1)<br>[32.7-41.8] | 641(34.8)<br>[30.7-39.2] | 705(40.5)<br>[35.9-45.3] | 694(38.8)<br>[34.2-43.7] | .32 | 2663(37.9)<br>[35.6-40.2] |
| Female | 707(35.3)<br>[30.9-39.9] | 791(34.8)<br>[31.4-38.4] | 838(39.3)<br>[35.4-43.3] | 821(37.7)<br>[33.1-42.7] | .33 | 3157(36.8)<br>[34.7-39.0] |
| *P* value | .37 | >.99 | .66 | .78 |  | .48 |
| **Age, y** |  |  |  |  |  |  |
| 20-39 | 205(17.4)<br>[14.6-20.6] | 227(20.1)<br>[16.1-24.7] | 241(21.2)<br>[17.1-26.0] | 204(21.7)<br>[17.1-27.3] | .35 | 877(20.1)<br>[18.1-22.4] |
| 40-59 | 484(40.9)<br>[35.2-47.0] | 533(37.4)<br>[34.5-40.5] | 556(46.3)<br>[39.6-53.1] | 509(41.6)<br>[36.8-46.5] | .08 | 2082(41.5)<br>[38.9-44.2] |
| ≥60 | 641(54.6)<br>[45.8-63.2] | 672(50.8)<br>[45.0-56.6] | 746(54.9)<br>[49.2-60.5] | 802(54.4)<br>[51.5-57.2] | .71 | 2861(53.7)<br>[50.7-56.7] |
| *P* value | <.001 | <.001 | <.001 | <.001 |  | <.001 |
| **Ethnicity** |  |  |  |  |  |  |
| Hispanic | 287(34.2) | 341(33.2) | 554(39.4) | 381(40.0) | .11 | 1563(36.9) |
|  | [28.4-40.4] | [28.5-38.4] | [33.8-45.3] | [36.2-43.9] |  | [34.4-39.4] |
| Non-Hispanic<br>White | 538(38.2)<br>[33.9-42.8] | 658(36.9)<br>[33.6-40.3] | 531(42.1)<br>[38.0-46.3] | 555(39.0)<br>[35.2-42.9] | .27 | 2282(39.3)<br>[37.1-41.0] |
| Non-Hispanic<br>Black | 373(33.5)<br>[30.0-37.2] | 310(33.2)<br>[30.0-36.5] | 306(33.4)<br>[28.5-38.7] | 338(33.9)<br>[30.2-37.8] | .99 | 1327(33.5)<br>[31.5-35.6] |
| Non-Hispanic<br>Asian | 103(21.8)<br>[16.7-28.0] | 95(19.3)<br>[15.4-23.8] | 91(22.9)<br>[18.3-28.3] | 156(31.2)<br>[27.5-35.3] | < .001 | 445(24.0)<br>[21.6-26.6] |
| Other | 29(35.0)<br>[15.3-61.6] | 28(30.0)<br>[19.3-43.4] | 61(49.8)<br>[38.7-61.0] | 85(41.9)<br>[28.7-56.3] | .14 | 203(41.2)<br>[33.7-49.1] |
| *P* value | <.001 | <.001 | <.001 | <.001 |  | <.001 |
| **Education level** |  |  |  |  |  |  |
| < 11th grade | 382(41.1)<br>[36.3-46.2] | 379(40.3)<br>[34.0-46.9] | 429(41.6)<br>[34.9-48.7] | 366(47.4)<br>[43.4-51.5] | .14 | 1556(42.3)<br>[39.4-45.3] |
| High School | 310(40.3)<br>[35.7-45.1] | 355(37.9)<br>[32.2-44.0] | 344(43.6)<br>[36.2-51.2] | 382(43.0)<br>[35.8-50.6] | .61 | 1391(41.4)<br>[38.0-44.9] |
| Some college | 390(38.6)<br>[31.8-45.8] | 456(38.7)<br>[33.9-43.7] | 482(42.3)<br>[37.3-47.5] | 483(38.8)<br>[33.2-44.7] | .70 | 1811(39.6)<br>[36.8-42.5] |
| College<br>graduate or<br>above | 247(28.3)<br>[22.0-35.6] | 241(25.8)<br>[22.9-28.8] | 287(34.1)<br>[29.2-39.3] | 281(29.5)<br>[24.8-37.4] | .05 | 1056(29.5)<br>[27.0-32.1] |
| *P* value | .003 | <.001 | .03 | <.001 |  | <.001 |
| **Poverty to income ratio** |  |  |  |  |  |  |
| <130% | 473(36.9)<br>[32.7-41.3] | 506(35.2)<br>[31.4-39.3] | 487(39.6)<br>[33.6-46.1] | 388(43.1)<br>[35.8-50.8] | .27 | 1854(38.4)<br>[35.7-41.2] |
| 130%-349% | 431(36.0)<br>[32.0-40.2] | 477(39.5)<br>[35.4-43.7] | 576(40.5)<br>[34.6-46.6] | 600(43.1)<br>[38.8-47.6] | .15 | 2084(39.9)<br>[37.5-42.2] |
| ≥350% | 319(35.0)<br>[28.3-42.3] | 341(31.9)<br>[27.4-36.8] | 331(40.0)<br>[34.7-45.4] | 345(33.5)<br>[28.4-39.0] | .16 | 1336(35.1)<br>[32.2-38.0] |
| *P* value | .86 | .03 | .99 | .006 |  | .02 |

### Trends in the prevalence of MetS in 2011-2018

The prevalence of MetS was 36.2% (95% CI, 32.3-40.3), 34.8% (95% CI, 32.3-37.4), 39.9% (95% CI, 36.6-43.2) and 38.3% (95% CI, 35.3-41.3) in 2011-2, 2013-4, 2015-6, 2017-8, respectively. The overall weighted prevalence of MetS remained stable across the four 2-year cycles (*P* for trend =.08). The percentage of MetS prevalence in non-Hispanic Asian increased from 21.8% (95% CI, 16.7-28.0) in 2011-2 to 31.2% (95% CI, 27.4-35.3) in 2017-8 [*P* for trend <.001; APC=14.6, (95% CI, 2.5-34.8)].

### Prevalence of MetS by socioeconomic status

High SES was associated with a low prevalence of MetS (Table 2). During 2011-2018, the weighted prevalence of MetS among college graduates was lower (29.5%; 95% CI, 27.0-32.1) than among those with some college (39.6%; 95% CI, 36.8-42.5) and those who were high school graduates or less [41.4% (95% CI, 38.0-44.9), 42.3% (95% CI, 39.4-45.3), respectively; *P* <.001]. The prevalence of MetS was lower in the household income ≥350% group (35.1%; 95% CI, 32.2-38.0) than the 130%-349% group and the <130% group [39.9% (95% CI, 37.5-42.2) and 38.4% (95% CI, 35.7-41.2), respectively; *P* =.02).

## Discussion

This is a report of the most up-to-date estimates of the prevalence of MetS in US adults. Our results showed no significant overall change in the prevalence of MetS since 2011. This conclusion is in line with a recent analysis of MetS prevalence in NHANES 2011-2016.^13^ As there might be a time lag between obesity and the development of diabetes and hypertension, it would be premature to conclude that the epidemic of obesity and diabetes would not result in a higher incidence of MetS in the future. Indeed, our analysis showed a rising trend in hyperglycemia, and so highlights the pressing need for measures to prevent diabetes in the US. On the basis of the 2018 census numbers for U.S. adults aged ≥20 years (∼247 million), we estimate that about 93 million people had the metabolic syndrome in 2018, which exceeded the prevalence of 50 million in 1990, and 64 million in 2000.^22^

We also investigated the prevalence of MetS in gender, age and ethnic subgroups. Older individuals have a higher prevalence of MetS due to the increase in the prevalence of hypertension and diabetes with age.^23, 24^ The increased overall prevalence of MetS was to a large extent driven by the increased prevalence in MetS in this age group. As the elderly population increases, the number of people with MetS is set to increase. A study showed that only 20.4% of older US adults had satisfactory health metrics.^7^ The increase in MetS and the associated increase in diabetes, hypertension and cardiovascular disease will increase the population disease burden in the coming years.^25^

The rise in the prevalence of MetS in non-Hispanic Asians in the US should not be overlooked. Asian Americans are the fastest-growing racial-ethnic minority in the United States. In 2018, Asian Americans comprised 6.5% of the U.S. population.^26^ Despite the seemingly low rate of obesity in Asian Americans, they are prone to MetS and type 2 diabetes for the same BMI as other ethnic groups.^27, 28^ Conversely, non-Hispanic blacks in the US have relatively higher lean mass and lower fat mass.^29^ The impact of high BMI on the development of diabetes and hypertension shows ethnic differences, with the greatest impact seen among Asians and the least impact among blacks.^30, 31^

We observed that the prevalence of MetS decreased with elevated educational attainment and income. People of low SES were more likely to have MetS. Possible explanations might include unhealthy lifestyle, diet habits, and poorer access to healthcare. Those with lower SES tend to have an unhealthy lifestyle, including smoking and alcohol consumption which could aggravate the potential risks of MetS.^32^ People with low income might prefer higher energy density food at less cost, which results in higher energy intake and increased risk of obesity.^33^ Obese and obesity are closely related to development of MetS. Low SES is also related to psychiatric illnesses which can also result in metabolic syndrome by inducing unhealthy behaviors such as alcohol consumption, smoking, poor diet, sleeping disorder and poor adherence to treatment.^34, 35^

MetS is associated with severe COVID-19.^7^ Nearly half of COVID-19 patients had a comorbidity such as hypertension and diabetes.^36^ Obesity confers a 2.4-fold higher odds of developing severe pneumonia in COVID-19.^37^ The high prevalence of MetS in the US and its increasing trend are worrying, since it may suggest that a substantial proportion of the US population is at risk of COVID-19 and its complications.

### Limitations

There are some limitations in this study that should be acknowledged. Firstly, the NHANES is a cross-sectional survey that collected data from different participants in each time interval failing to provide longitudinal follow-up data. Secondly, NHANES used a recall questionnaire for some variables, especially the antihypertension and anti-diabetes medications, prone to recall and response bias. Thirdly, the number of participants, especially with triglyceride and high-density lipoprotein cholesterol, is relatively small.

## Conclusions

The prevalence of MetS in the US has not increased significantly in 2011-8. It was 36.2% (95% CI, 32.3-40.3) in 2011-2 and 38.3% (95% CI, 35.3-41.3) in 2017-8. This may be a reflection of the type 2 diabetes epidemic in the US. Lifestyle modification including healthy diet and regular physical activity would help to reduce the risk of developing MetS. This is especially important in Asian Americans and people of low SES. The early recognition of MetS, the prompt modifications of lifestyle may forestall the late complications of MetS including cardiovascular disease.

## Data Availability

All the data including demographics, examination, questionnaire, and laboratory data covering the period 2011-2018 were extracted.19. National Center for Health Statistics. National Health and Nutrition Examination Survey. Available at: http://www.cdc.gov/nchs/nhanes.htm.

## Author Contribution

B Cheung has full access to all of the data in the study and takes responsibility for the integrity of the data and the accuracy of the data analysis.

Concept and design: all authors.

Acquisition, analysis, or interpretation of data: Liang, B Cheung

Drafting of the manuscript: Liang, B Cheung

Critical revision of the manuscript for important intellectual content: all authors

Statistical analysis: Liang, Or, Tsoi

Administrative, technical, or material support: B Cheung

Supervision: B Cheung

## Acknowledgment

BMY Cheung is the Sun Chieh Yeh Heart Foundation Professor in Cardiovascular Therapeutics and receives funding from the foundation.

## References

1. Alberti KG, Zimmet P, Shaw J; IDF Epidemiology Task Force Consensus Group. The metabolic syndrome--a new worldwide definition. Lancet. 2005;366(9491):1059–1062.

2. Third Report of the National Cholesterol Education Program (NCEP) Expert Panel on Detection, Evaluation, and Treatment of High Blood Cholesterol in Adults (Adult Treatment Panel III) final report. Circulation. 2002;106(25):3143–3421.

3. Cheung BM, Wat NM, Man YB, et al. Development of diabetes in Chinese with the metabolic syndrome: a 6-year prospective study. Diabetes Care. 2007;30(6):1430–1436.

4. Cheung BM, Wat NM, Man YB, et al. Relationship between the metabolic syndrome and the development of hypertension in the Hong Kong Cardiovascular Risk Factor Prevalence Study-2 (CRISPS2). Am J Hypertens. 2008;21(1):17–22.

5. Cheung BM. The cardiovascular continuum in Asia--a new paradigm for the metabolic syndrome. J Cardiovasc Pharmacol. 2005;46(2):125–129.

6. Yanai H. Metabolic Syndrome and COVID-19. Cardiol Res. 2020;11(6):360–365.

7. Li HL, Cheung BMY. The Proportion of Adult Americans at Risk of Severe COVID-19 Illness. J Gen Intern Med. 2021;36(1):259–261.

8. Thomas GN, Schooling CM, McGhee SM, et al. Metabolic syndrome increases all-cause and vascular mortality: the Hong Kong Cardiovascular Risk Factor Study. Clin Endocrinol (Oxf). 2007;66(5):666–671.

9. Cheng YJ, Imperatore G, Geiss LS, et al. Secular changes in the age-specific prevalence of diabetes among U.S. adults: 1988-2010. Diabetes Care. 2013;36(9):2690–2696.

10. Després JP, Lemieux I. Abdominal obesity and metabolic syndrome. Nature. 2006;444(7121):881–887.

11. Shin D, Kongpakpaisarn K, Bohra C. Trends in the prevalence of metabolic syndrome and its components in the United States 2007-2014. Int J Cardiol. 2018;259:216-219.

12. Falkner B, Cossrow ND. Prevalence of metabolic syndrome and obesity-associated hypertension in the racial ethnic minorities of the United States. Curr Hypertens Rep. 2014;16(7):449.

13. Hirode G, Wong RJ. Trends in the Prevalence of Metabolic Syndrome in the United States, 2011-2016. JAMA. 2020;323(24):2526–2528.

14. Mozumdar A, Liguori G. Persistent increase of prevalence of metabolic syndrome among U.S. adults: NHANES III to NHANES 1999-2006. Diabetes Care. 2011;34(1):216–219.

15. Aguilar M, Bhuket T, Torres S, Liu B, Wong RJ. Prevalence of the metabolic syndrome in the United States, 2003-2012. JAMA. 2015;313(19):1973–1974.

16. Grundy SM. Pre-diabetes, metabolic syndrome, and cardiovascular risk. J Am Coll Cardiol. 2012;59(7):635–643.

17. Adler NE, Ostrove JM. Socioeconomic status and health: what we know and what we don’t. Ann N Y Acad Sci. 1999;896:3-15.

18. Kanjilal S, Gregg EW, Cheng YJ, et al. Socioeconomic status and trends in disparities in 4 major risk factors for cardiovascular disease among US adults, 1971-2002. Arch Intern Med. 2006;166(21):2348–2355.

19. National Center for Health Statistics. National Health and Nutrition Examination Survey. Available at: http://www.cdc.gov/nchs/nhanes.htm. Accessed January 1, 2021.

20. Grundy SM, Cleeman JI, Daniels SR, et al. Diagnosis and management of the metabolic syndrome: an American Heart Association/National Heart, Lung, and Blood Institute Scientific Statement. Circulation. 2005;112(17):2735–2752.

21. National Center for Health Statistics: NHANES: Response Rates and Population Totals. https://www.n.cdc.gov/nchs/nhanes/ResponseRates.aspx#response-rates. Accessed December 21, 2020.

22. Ford ES, Giles WH, Mokdad AH. Increasing prevalence of the metabolic syndrome among u.s. Adults. Diabetes Care. 2004;27(10):2444–2449.

23. Fryar CD, Ostchega Y, Hales CM, Zhang G, Kruszon-Moran D. Hypertension Prevalence and Control Among Adults: United States, 2015-2016. NCHS Data Brief. 2017;(289):1–8.

24. Corriere M, Rooparinesingh N, Kalyani RR. Epidemiology of diabetes and diabetes complications in the elderly: an emerging public health burden. Curr Diab Rep. 2013;13(6):805–813.

25. Ferrucci L, Fabbri E. Inflammageing: chronic inflammation in ageing, cardiovascular disease, and frailty. Nat Rev Cardiol. 2018;15(9):505–522.

26. Dhooper SS. Health care needs of foreign-born Asian Americans: an overview. Health Soc Work. 2003;28(1):63–73.

27. Palaniappan LP, Wong EC, Shin JJ, Fortmann SP, Lauderdale DS. Asian Americans have greater prevalence of metabolic syndrome despite lower body mass index. Int J Obes (Lond). 2011;35(3):393–400. doi:10.1038/ijo.2010.152

28. Qian Y, Lin Y, Zhang T, et al. The characteristics of impaired fasting glucose associated with obesity and dyslipidaemia in a Chinese population. BMC Public Health. 2010;10:139. Published 2010 Mar 17.

29. Schutte JE, Townsend EJ, Hugg J, Shoup RF, Malina RM, Blomqvist CG. Density of lean body mass is greater in blacks than in whites. J Appl Physiol Respir Environ Exerc Physiol. 1984;56(6):1647–1649.

30. Spanakis EK, Golden SH. Race/ethnic difference in diabetes and diabetic complications. Curr Diab Rep. 2013;13(6):814–823.

31. Colin Bell A, Adair LS, Popkin BM. Ethnic differences in the association between body mass index and hypertension. Am J Epidemiol. 2002;155(4):346–353.

32. Zhan Y, Yu J, Chen R, et al. Socioeconomic status and metabolic syndrome in the general population of China: a cross-sectional study. BMC Public Health. 2012;12:921.

33. Lipsky LM. Are energy-dense foods really cheaper? Reexamining the relation between food price and energy density. Am J Clin Nutr. 2009;90(5):1397–1401.

34. Lorant V, Deliège D, Eaton W, Robert A, Philippot P, Ansseau M. Socioeconomic inequalities in depression: a meta-analysis. Am J Epidemiol. 2003;157(2):98–112.

35. Foley DL, Morley KI, Madden PA, Heath AC, Whitfield JB, Martin NG. Major depression and the metabolic syndrome. Twin Res Hum Genet. 2010;13(4):347–358.

36. Zhou F, Yu T, Du R, et al. Clinical course and risk factors for mortality of adult inpatients with COVID-19 in Wuhan, China: a retrospective cohort study. Lancet. 2020;395(10229):1054–1062.

37. Cai Q, Chen F, Wang T, et al. Obesity and COVID-19 Severity in a Designated Hospital in Shenzhen, China. Diabetes Care. 2020;43(7):1392–1398.

